# Predicting new onset thought disorder in early adolescence with optimized deep learning implicates environmental-putamen interactions

**DOI:** 10.1101/2023.10.23.23297438

**Authors:** Nina de Lacy, Michael J. Ramshaw

## Abstract

**Background:** Thought disorder (TD) is a sensitive and specific marker of risk for schizophrenia onset. Specifying factors that predict TD onset in adolescence is important to early identification of youth at risk. However, there is a paucity of studies prospectively predicting TD onset in unstratified youth populations.

**Study Design:** We used deep learning optimized with artificial intelligence (AI) to analyze 5,777 multimodal features obtained at 9-10 years from youth and their parents in the ABCD study, including 5,014 neural metrics, to prospectively predict new onset TD cases at 11-12 years. The design was replicated for all prevailing TD cases at 11-12 years.

**Study Results:** Optimizing performance with AI, we were able to achieve 92% accuracy and F1 and 0.96 AUROC in prospectively predicting the onset of TD in early adolescence. Structural differences in the left putamen, sleep disturbances and the level of parental externalizing behaviors were specific predictors of new onset TD at 11-12 yrs, interacting with low youth prosociality, the total parental behavioral problems and parent-child conflict and whether the youth had already come to clinical attention. More important predictors showed greater inter-individual variability.

**Conclusions:** This study provides robust person-level, multivariable signatures of early adolescent TD which suggest that structural differences in the left putamen in late childhood are a candidate biomarker that interacts with psychosocial stressors to increase risk for TD onset. Our work also suggests that interventions to promote improved sleep and lessen parent-child psychosocial stressors are worthy of further exploration to modulate risk for TD onset.

## INTRODUCTION

Thought disorder (TD) is a disruption in the flow of thought and language processing inferred from disorganized speech and a cardinal symptom of youth and adult schizophrenia. (1-4) A distinct construct within the psychotic phenotype, it is a sensitive and specific indicator of schizophrenia, is associated with the most severe illness and can persist after positive symptom resolution where this persistence is a marker of poor outcomes. (1, 5-11) TD also frequently exists prior to frank schizophrenia onset and was first identified as a high-risk prodromal marker in the 1980s. (12) Many subsequent studies over the last two decades have similarly demonstrated that TD and conceptual disorganization are strong predictors of later transition to frank psychosis. (13-17) Importantly, re-analysis of data from the large (n=764) North American Prodrome Longitudinal Study-2 (NAPLS-2) further indicates that thought disorganization distinguishes among different subgroups of prodromal patients and is a stronger predictor of onset than perceptual abnormalities, (18, 19) a result replicated in an independent external cohort. (20) TD shows familial aggregation, (21) being present in unaffected relatives of people with schizophrenia (22-27) - in contradistinction to bipolar disorder (28) - as well as unaffected twins in discordant twin pairs. (29) It has also been detected in people with schizotypal personality disorder. (1) Collectively, these phenomena suggest it may be an endophenotype or marker for genetic risk for schizophrenia. (21, 30, 31)

Prodromal research in adolescents and young adults has most often followed individuals at elevated risk for schizophrenia in longitudinal designs and retrospectively quantified associations between their evolving symptom profiles and conversion to frank psychosis. For example, teenagers in outpatient care with extant subclinical symptoms or at high familial risk for schizophrenia. The resultant improved delineation of the prodromal phase has supported the development of early intervention programs, which have shown promise. The latter are particularly important since the duration of untreated psychosis (DUP) is well-associated with poorer long-term prognoses (32-37) and reductions in neurocognitive abilities, (38) albeit some studies have questioned these findings (39). A recent large-scale meta-analysis concluded that effect sizes were clinically meaningful, with “a DUP of four weeks predicting >20% more severe symptoms at follow-up relative to a DUP of one week.” (40) Thus, identifying risk factors, behavioral indicators and/or candidate biomarkers that predispose individuals to develop schizophrenia is important to reduce morbidity. Further, the relative paucity of new drug treatments for psychosis in recent decades has fueled interest in modifiable factors that may be addressed to mitigate outcomes. Studies in identical twins indicate that pairwise concordance for schizophrenia is only ∼50%. (41) Thus, environmental factors would appear to play a substantial role in relative risk, likely interacting with aberrant microglial function leading to loss of dendritic spines in select neuronal populations, evident at the macroscale in gray matter volume loss and brain network organizational differences at disease onset associated with cognitive deficits. (42-45)

Evidence from a number of avenues suggests that the pathogenesis of schizophrenia is associated with altered neurodevelopment from early life, including *in utero* exposures, obstetric complications and neurocognitive differences. (46-50) Other environmental factors in childhood and adolescence that have been associated with schizophrenia risk include urban living, migration (first generation), cannabis use and adverse or threatening events. (51, 52) Extant research suggests that environmental stressors act upon cortical and subcortical stress-response circuitry to sensitize and dysregulate the striatal dopamine system. (53-58) For example, adverse events and unstable family environments in childhood have been linked to elevated striatal dopamine function in adulthood in youth at ultra-high risk for schizophrenia. (54)

Given the importance of identifying youth at risk for schizophrenia to minimize DUP and the position of TD as the strongest predictor of onset, understanding which factors predict the onset of TD itself is an important question. (59) While direct and/or specific causation cannot yet be inferred between the development of TD and later development of schizophrenia, identifying high-value risk factors, behavioral indicators and/or candidate biomarkers of TD onset might concomitantly increase understanding of risk, pathogenesis and trajectories of schizophrenia onset and support early intervention efforts. Unfortunately, there is a paucity of studies predicting TD onset as opposed to examining the prognostic value of TD in predicting schizophrenia onset. A substantial barrier has been the lack of appropriate multi-domain data in large participant samples of youth with appropriate TD assessment. Outside the US, national registries or school system data may be available offering large sample sizes (*n*>10,000) but these typically lack physiologic, psychometric and TD information. (60-63) However, in recent years data has started to appear from the landmark Adolescent Brain Cognitive Development (ABCD) study. Sponsored by NIH, this is the largest long-term study of child and adolescent brain development and health in the US. ABCD recruited ∼11,800 children age 9-10 years (yrs) and is following this cohort for at least a decade with a comprehensive battery of psychosocial, biological, cognitive, cultural and environmental repeated assessments including multiple modalities of neuroimaging conducted every 2 yrs. TD is assessed by ABCD, offering an invaluable opportunity to study the onset of TD in early and mid-adolescence, which are critical sensitive periods preceding the typical prodromal period.

In the present study, we aimed to prospectively predict the onset of TD in early adolescence. Specifically, we predicted the onset of TD in early adolescence at 11-12 yrs using assessments collected in late childhood at 9-10 yrs in the ABCD cohort. Since the existing literature is scant, we threw as wide as possible a net and analyzed 5,777 multimodal candidate predictors encompassing demographic, developmental, academic, psychosocial, biological, cognitive, cultural and environmental measures and metrics drawn from multiple brain imaging modalities (white and gray matter brain structure and volumetrics; cortical and subcortical connectivity and 3 cognitive tasks). Concomitantly, we employed deep learning (DL) with artificial neural networks to make prospective predictions of case onset and provide multivariable signatures of TD valid at the person-level. While DL offers powerful predictive capability and is resistant to multicollinearity, its application to translational aims can be limited by the relative difficulty of tuning these models and their tendency to act as ‘black box’ estimators where the features used to make predictions are not interpretable. Accordingly, we enhanced DL performance with Integrated Evolutionary Learning (IEL), an AI-based form of computational intelligence, to jointly optimize across the hyperparameters, learn the most important final predictors of TD and render interpretable predictions. This is a massively parallel approach where ∼40,000 model fits are pursued to train and optimize models before testing final, optimized models in a holdout, unseen data partition. All results presented are from testing in this holdout, unseen data.

## MATERIALS AND METHODS

### Terminology and definitions

This manuscript uses machine learning (ML) conventions throughout. (64-66) ‘Prediction’ means predicting the quantitative value of a target variable by analyzing patterns in features (input data). Specifically, we also use it here to mean prospective prediction i.e. predicting an outcome in the future from *ex ante* features. The set of all input data is referred to as containing ‘features’ (candidate predictors) and those features contributing to final, optimized models (presented in **Results)** as ‘predictors’. The set of observations (features and targets) used to train and validate models is referred to as the ‘training set’ and the unseen holdout set of observations is termed the ‘test set’. ‘Generalizability’ is defined as the ability of a trained model to adapt to new, previously unseen data drawn from the same distribution i.e. model fit in the test set. ‘Precision’ refers to the fraction of positive predictions that were correct; ‘Recall’ to the proportion of true positives that were correctly predicted; ‘Accuracy’ to the number of correct predictions as a fraction of total predictions; and ‘F1’ to the harmonic mean of precision and recall (i.e. how many times a correct prediction was obtained across the target). Receiver Operating Characteristic curves (ROC Curves) are provided that quantify classification performance at different classification thresholds plotting true positive versus false positive rates, where the Area Under the Receiver Operating Characteristic Curve (AUROC) is defined as the two-dimensional area under the ROC curve from (0,0) to (1,1).

### Data and data collection in the ABCD study

ABCD is an epidemiologically informed prospective cohort study that recruited 11,880 children (52% male; 48% female) at ages 9-10 years (108-120 months) via 21 sites across the United States. It is oversampled for twin pairs (*n*=800) and non-twin siblings may also be enrolled. ABCD study data is available to qualified researchers from the National Institute of Mental Health Data Archive (NDA) and is released periodically with no embargo. This study uses data from release 4.0, which includes data up to 42-month follow-up. Further detail on the study design, the construction of the participant sample and recruitment procedures may be found in Jernigan et al; Garavan et al; and Volkow et al. (67-69)

ABCD collects a rich set of repeated assessments about youth and their families. Broadly, phenotypic assessments are performed annually and multimodal neuroimaging every 2 years. The phenotypic and substance abuse assessment protocol is covered in detail in Barch et al and Lisdahl et al, respectively. (70, 71) In brief, data collected at 9-10 yrs includes measures of demographics, physical and mental health, substance use, cognition, culture and environment that are collected for children and their parents and biospecimen collection for DNA, pubertal hormone levels, substance use metabolites (hair) and substance and environmental toxin exposure (baby teeth) from youth. These are supplemented with geocoded environmental variables. A summary description of features used here may be inspected in **Supplementary Table 1**. Brain imaging protocols comprise optimized 3D T1; 3D T2; Diffusion Tensor Imaging; Resting state functional MRI (rsfMRI); and 3 task MRI (tfMRI) protocols that are harmonized to be compatible across acquisition sites. The 3 cognitive tfMRI tasks are the Monetary Incentive Delay and Stop Signal tasks and an emotional version of the n-back task which collectively measure reward processing, motivation, impulsivity and emotion regulation, impulse control and working memory. ABCD provides fully-processed metrics from each of these imaging types via the NDA. Full details of the brain imaging protocol may be inspected in Casey et al and the pre-processing and analytic pipeline used to generate the neural metrics used here in Hagler et al. (72, 73) We used all available processed metrics from diffusion fullshell, cortical and subcortical Gordon correlations (rsfMRI), gray and white matter structural and volumetric processing and each cognitive tasks. Head motion statistics for each modality and acquisition site were also included in the feature set. For certain neuroimaging modalities, multiple scans were attempted or completed and here we use metrics obtained from the first scan since these were available for the largest number of participants.

The present study was deemed not human subjects research by the University of Utah Institutional Review Board.

### Study inclusion criteria and sample partitioning for machine learning

Inclusion criteria for the present study were a) participants enrolled in the study at baseline who were still enrolled at 2-year follow-up (*n*=8,085) who had b) complete data passing ABCD quality control that was available for all neural metric types (*n*=6,178) and were c) singleton participants (i.e. unrelated to any other youth participant in the study (*n*=5,356)). If a youth had a sibling also enrolled in the study, we selected the oldest sibling for inclusion in our study. Demographic characteristics of our sample at age 9-10 yrs are presented in **Table 1**.

**Table 1:** Demographic characteristics of participant sample at age 9-10 years. Sex refers to natal sex as reported on the youth’s original birth certificate. Gender refers to the youth’s reported gender identification. Race and ethnicity refer to parent report of youth’s race or ethnicity. More than one race or ethnicity identification may be selected and therefore these percentages may sum to >100%.

| Characteristic | Number | Percent |
| --- | --- | --- |
| **Sex** |  |  |
| Male | 2,771 | 51.7% |
| Female | 2,584 | 48.3 |
| **Reported Gender** |  |  |
| Male | 2,768 | 51.7% |
| Female | 2,577 | 48.1 |
| Gender non-conforming | 7 | 0.1 |
| Don't know/didn't answer | 4 | 0.1 |
| **Reported Race** |  |  |
| Black/African American | 873 | 16.3% |
| Asian | 353 | 6.6 |
| White | 4,236 | 79.1 |
| Native American/Alaska Native | 187 | 3.5 |
| Other | 334 | 6.2 |
| **Reported Ethnicity** |  |  |
| Hispanic/Latino/Latinx | 1,070 | 20.0% |
| Non-Hispanic | 4,224 | 78.9 |
| Not indicated | 62 | 1.2 |

Physiologic and cognitive characteristics of our participant sample at 9-10 yrs may be viewed in **Table 2**.

**Table 2:**
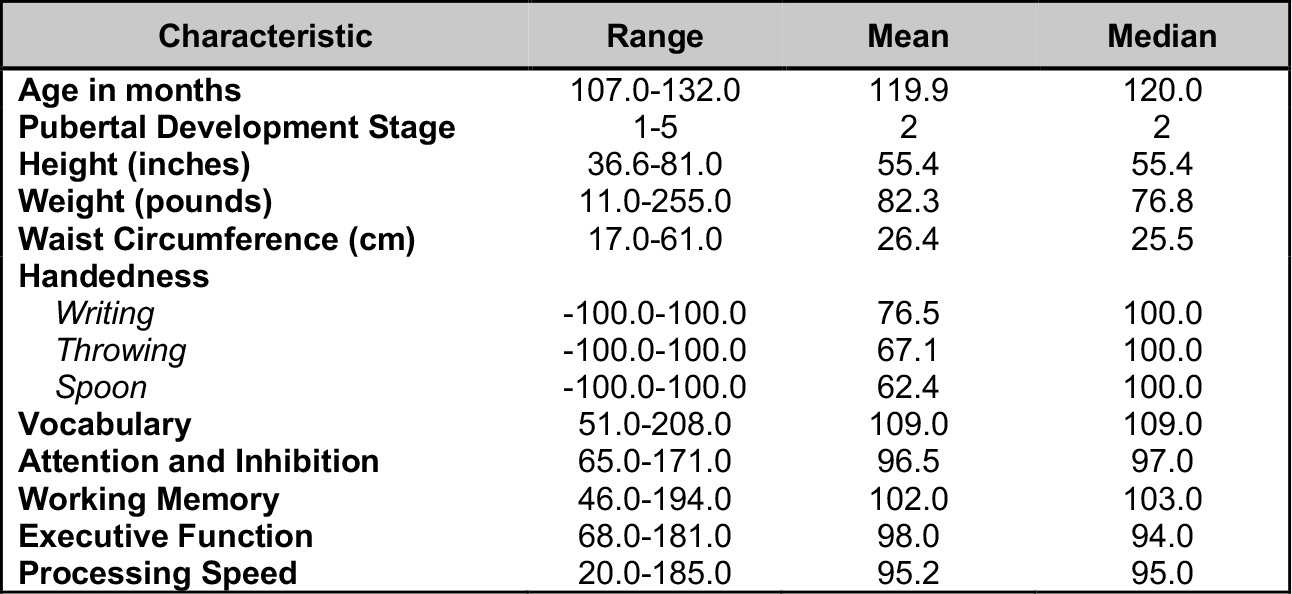
Physiologic and cognitive characteristics of participant sample at age 9-10 years. Characteristics of the study sample at 9-10 yrs. Pubertal development was measured with the Pubertal Development Scale (adapted from the Petersen scale) in a sex-specific manner. Height was measured twice with the average of these values reported here. We note a range of 11.0-255.0 pounds for weight which is the range present in the original ABCD data. Handedness was assessed with the Edinburgh Handedness Inventory. Age-corrected cognitive scores were assessed with the NIH Toolbox. Vocabulary was measured with the Picture Vocabulary test; Attention and inhibition with the Flanker Inhibitory Control & Attention Task; Executive Function with the Dimensional Change Card Sort Test; and Processing Speed with the Pattern Comparison Processing Speed Test.

| Characteristic | Range | Mean | Median |
| --- | --- | --- | --- |
| Age in months | 107.0-132.0 | 119.9 | 120.0 |
| Pubertal Development Stage | 1-5 | 2 | 2 |
| Height (inches) | 36.6-81.0 | 55.4 | 55.4 |
| Weight (pounds) | 11.0-255.0 | 82.3 | 76.8 |
| Waist Circumference (cm) | 17.0-61.0 | 26.4 | 25.5 |
| Handedness |  |  |  |
| Writing | -100.0-100.0 | 76.5 | 100.0 |
| Throwing | -100.0-100.0 | 67.1 | 100.0 |
| Spoon | -100.0-100.0 | 62.4 | 100.0 |
| Vocabulary | 51.0-208.0 | 109.0 | 109.0 |
| Attention and Inhibition | 65.0-171.0 | 96.5 | 97.0 |
| Working Memory | 46.0-194.0 | 102.0 | 103.0 |
| Executive Function | 68.0-181.0 | 98.0 | 94.0 |
| Processing Speed | 20.0-185.0 | 95.2 | 95.0 |

The resulting group of 5,356 participants was then randomly partitioned into a training set comprising 70% of the sample (*n*=3,749) and a holdout, unseen test set comprising 30% of the sample (*n*=1,607). This partitioning was performed prior to pre-processing either features or the predictive target of TD to minimize bias.

### Preparation of predictive targets

The present study uses predictive targets for TD derived from the Child Behavior Checklist (CBCL) for youth ages 4-18 yrs. The CBCL is a standardized instrument in widespread clinical and research use that forms part of the Achenbach System of Empirically Based Assessment (ASEBA) that is “designed to facilitate assessment, intervention planning and outcome evaluation among school, mental health, medical and social service practitioners who deal with maladaptive behavior in children, adolescents and young adults.” (74) Parents rate their child on a 0-1-2 scale on 118 specific problem items that are aggregated into raw, T and percentile scores for syndrome subscales derived from principal components analysis of data from 4,455 children referred for mental health services. The CBCL is normed in a sex/gender-specific manner on a U.S. nationally representative sample of 2,368 youth ages 4-18 that takes into account differences in problem scores for males versus females. It exhibits excellent test-retest reliability of 0.82-0.96 for the syndrome scales with an average *r* of 0.89 across all scales. Content and criterion validity is strong with referred versus non-referred children scoring higher on 113/188 problem items and significantly higher on all problem scales, respectively. In the present analysis, the syndrome subscale for Thought Problems was used to assess TD. The CBCL Thought Problem Scale contains 15 items assessing obsessive thoughts, self-harm, hallucinations, nervous twitching, picking parts of body, playing with own sex parts in public and/or too much, compulsions, less need for sleep, storing too many items, strange behavior or ideas, sleep walking or talking, and trouble sleeping. Prior work has validated the CBCL Thought Problem subscale for detecting psychotic symptoms in children. (75) To form binary classification targets, CBCL T-scores were thresholded for each participant using cutpoints established by ASEBA for clinical practice. Specifically, T scores of 65-69 (95^th^ to 98^th^ percentile) are considered ‘borderline clinical’ and scores of >70 in the ‘clinical range.’ Accordingly, every individual with a T score ≥ 65 was deemed a ‘case’ [1] and every individual with a score <65 as a ‘not case’ [0] to discretize their T score. This process was performed separately in the training and test sets.

### Construction of participant case samples

We formed two different participant samples (**Figure 1**). In both, a TD case was defined as in **Preparation of predictive targets**. The first contained only new onset cases of TD defined as a youth age 11-12 yrs meeting criteria for TD who did not meet criteria at 9-10 yrs. The second sample contained all prevailing TD cases at 11-12 yrs. We constructed a balanced sample of controls for each of these samples from the eligible study population (see: **Baseline inclusion criteria and sample partitioning for machine learning**) comprising youth that were matched with cases by age and natal sex and had the lowest possible scores on the CBCL TD subscale. Both the new onset (*n*=288) and prevailing case (*n*=526) samples in the training partitions comprised >200 participants, a recommended threshold for robust ML classification.

**Figure 2:**
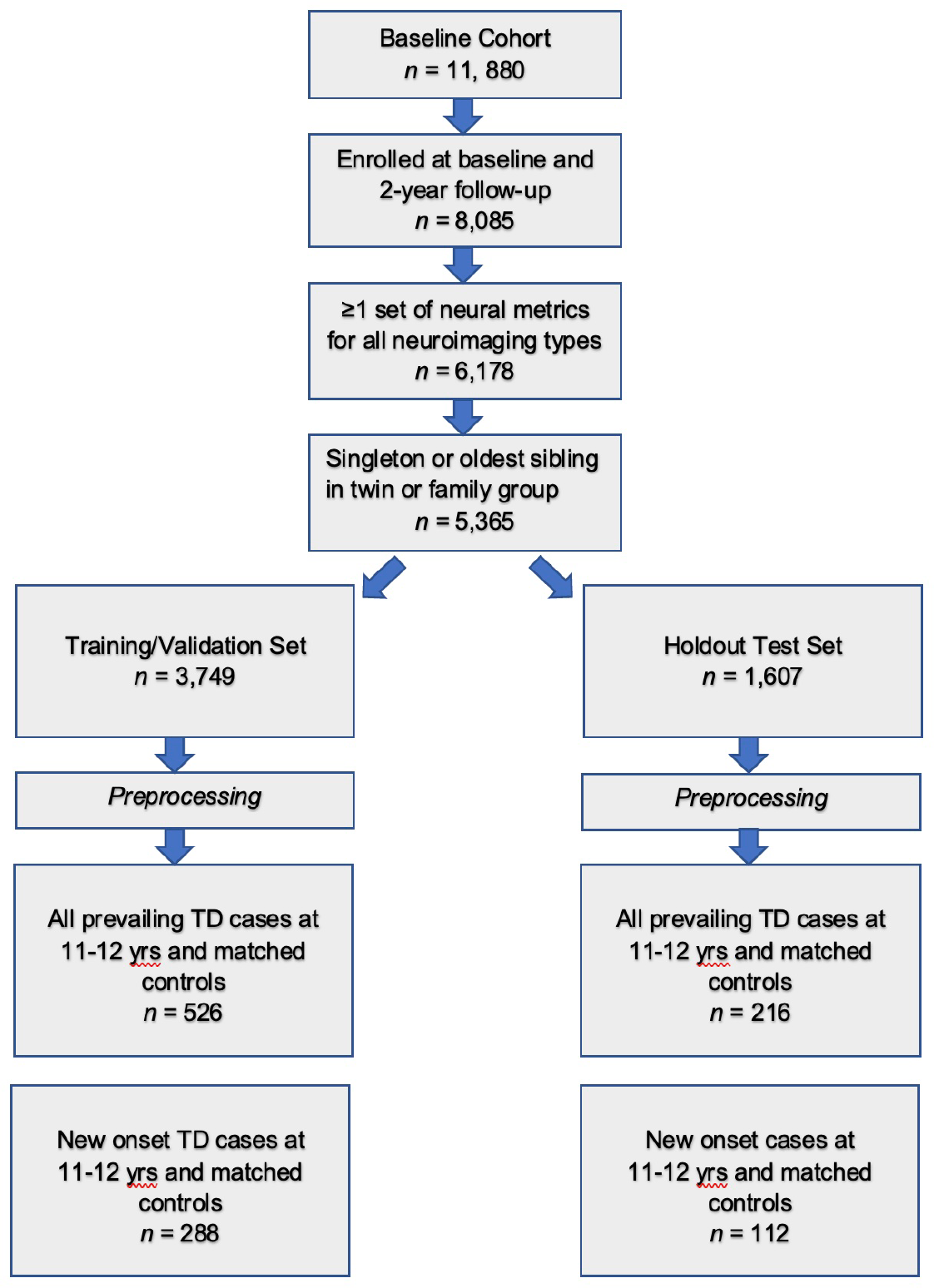
Construction of participant case samples. Steps used to form the study sample are displayed. After inclusion criteria were applied, the sample was randomly partitioned into training and test sets. Subsequently, samples for each experiment were formed as described in **Preparation of predictive targets** and **Construction of participant case samples**.

### Feature preparation

Our feature set included the majority of available phenotypic and environmental variables available from the ABCD study and all available neural metrics with the exception of temporal variance measures. Of note, metrics or instruments that directly quantified mental health symptoms were excluded (e.g. the Prodromal Psychosis Scale) to avoid redundancy and bias. For continuous features, subscale or total scores were used where available. Feature pre-processing was subsequently performed separately in the training and test sets to ready input data for ML. Firstly, features with >35% missing values were discarded: prior work shows that good results may be obtained with ML methods with imputation up to 50% missing data. (76) Nominal variables were one-hot encoded to transform them into discrete variables. Continuous variables were trimmed to [mean +/- 3] standard deviations to remove outliers and scaled in the interval [0,1] with the MinMaxScaler. Missing values were imputed using non-negative matrix factorization (NNMF). NNMF is a mathematically-proven imputation method that minimizes the cost function of missing data rather than assuming zero values that is effective at capturing both global and local structure in the data. It is particularly suitable for analyses using large-scale multimodal data since it has been demonstrated to perform well regardless of the underlying pattern of missingness. (77-79) **Supplementary Table 2** shows the number and percentage of observations which were trimmed and filled with NNMF for the training and test sets, respectively. After imputation with NNMF, any variables originating from phenotypic assessments lacking summary scores were reduced to single continuous, summary metrics using feature agglomeration. After pre-processing measures, a final set of 763 phenotypic and environmental features was obtained. Neural metrics (*n*=5,014) were processed by the ABCD study team and were therefore not pre-processed with the exception of scaling, again performed separately in the training and test partitions. There were no missing neural features. The final combined feature set including neural, phenotypic, environmental, head motion and site features comprised 5,777 features.

### Overview of predictive analytic pipeline

Deep learning was used to prospectively predict future cases of TD at 11-12 yrs using data collected at 9-10 yrs from youth and their parents and environmental data in two scenarios. Firstly, new onset cases at 11-12 yrs and secondly all prevailing cases at 11-12 yrs. All predictive models were implemented with *k*-fold cross-validation during the training process where the latter was supervised by an AI meta-learning algorithm that jointly performed feature selection and optimized across deep learning hyperparameters in an automated manner, pursuing ∼40,000 model fits for each experiment. Model training was terminated based on the Bayes Information Criterion (BIC), an information theoretic metric. The ability of final, optimized models to generalize was tested in the unseen test partition and performance statistics of AUROC, accuracy, precision and recall were computed and ROC curves generated based on this testing. The importance of predictors to making case predictions quantified with Shapley Additive Explanations (SHAP).

### Feature Filtering

We performed feature filtering prior to beginning model training in order to determine which of the 5,777 features exhibited a non-zero relationship with the predictive target (TD cases) and thereby reduce the number of features entering deep learning in a principled manner. This was accomplished in two stages. Firstly, a simple filtering process was performed in which χ^2^ (categorical features) and ANOVA (continuous features) statistics and the mutual information metric (all features) were computed among each feature and the target (TD cases) represented by a categorical vector in [0,1]. Any feature with a non-zero relationship (either positive or negative) with the target was retained. Secondly, feature selection was performed on these filtered feature subsets using the Least Absolute Shrinkage and Selection Operator (LASSO) algorithm. The LASSO is a well-established regularization technique that efficiently selects a reduced set of features by forcing certain regression coefficients to zero. We implemented the LASSO within AI meta-learning algorithm Integrated Evolutionary Learning to tune the α hyperparameter in the same manner as described below in **Integrated Evolutionary Learning for deep learning optimization**. This hyperparameter (commonly called the α) instantiates the amount of penalization (shrinkage) that will be imposed on features. The number of features **Table 2** remaining after each filtering step may be viewed in **Table 2**. Specific features selected in the LASSO process and resulting univariate coefficients between each of these features and the TD targets for each participant sample (new onset cases and all prevailing cases at 11-12 yrs) may be viewed in **Supplementary Table 3a** and **3b**, respectively.

### Deep learning with artificial neural networks

We used deep learning with artificial neural networks to prospectively predict TD cases. We trained artificial neural networks using the AdamW algorithm with 3 layers, 300 neurons per layer, early stopping (patience = 3, metric = validation loss) and the Relu activation function. The last output layer contained a conventional softmax function. Learning parameters (**Table 3**) were tuned with IEL as detailed below. Deep learning models were encoded with TensorFlow embedded in custom Python code.

**Table 3:**
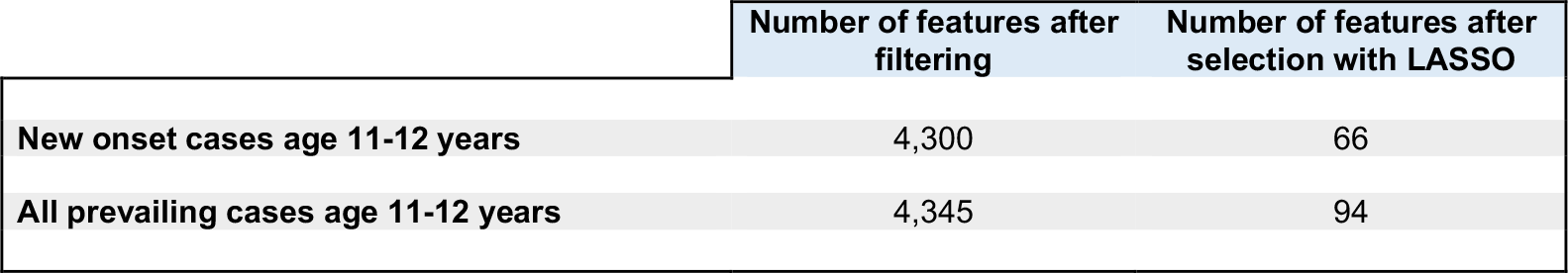
Feature sets after feature filtering. The total baseline set of 5,777 features was reduced in a two-step process, with simple feature filtering followed by regularization with the LASSO algorithm. The number of features remaining after each step is shown for new onset and all prevailing TD cases at 11-12 yrs. Detailed tables showing the univariate coefficients between each feature selected by the LASSO and TD target for each participant sample may be viewed in **Supplementary Tables 3a** and **3b**.

|  | Number of features after filtering | Number of features after selection with LASSO |
| --- | --- | --- |
| New onset cases age 11-12 years | 4,300 | 66 |
| All prevailing cases age 11-12 years | 4,345 | 94 |

### Integrated Evolutionary Learning for optimization across hyperparameters and feature selection

Many ML algorithms have hyperparameters that control learning that require ‘tuning’ during the training process and can have a substantial effect on model fit, performance and results. Typically, tuning is performed via local heuristics and ≤50 model fits are pursued, introducing the possibility of bias and potentially limiting the solution space. (80-82) Moreover, model fitting with large numbers of features introduces the challenge of avoiding overfitting while still retaining translational interpretability. To address these challenges, we previously developed and here applied an AI technique called Integrated Evolutionary Learning which can improve the performance of ML predictive algorithms in comparable tabular data by up to 20-25% versus the use of default model hyperparameters and conventional designs. (83) IEL is a form of computational intelligence based on evolutionary algorithms, which instantiate the concepts of biological evolutionary selection in computer code. IEL jointly optimizes across deep learning hyperparameters (**Table 4**) and selects high-value predictors by adaptively breeding models over hundreds of learning generations by selecting for improvements in a fitness function, the BIC.

**Table 4:** Hyperparameter settings optimized with Integrated Evolutionary Learning. Optimization across the hyperparameters of learning rate, Beta 1 and Beta 2 was conducted for deep learning with artificial neural networks within the ranges shown.

| Hyperparameters | Range | Mutation Shift |
| --- | --- | --- |
| Learning rate | 0.00001-0.01 | 0.0001 |
| Beta 1 | 0.9-0.999 | 0.001 |
| Beta 2 | 0.9-0.999 | 0.001 |

At the start of training, IEL initialized a first generation of 100 models with randomized hyperparameter values and feature sets, collectively ‘chromosomes’. Hyperparameter settings were subsequently recombined, mutated or eliminated over successive generations. In recombination, ‘parent’ hyperparameters are arithmetically averaged to form ‘children’. In mutation, hyperparameter settings are shifted with the range of possible values shown in **Table 4**. After the first generation of 100 models were trained, the BIC was computed for each solution. Of the 80 best models (lowest BIC), 40 were recombined by averaging the hyperparameter setting after a pivot point at the midpoint to produce 20 ‘child’ models. 20 were mutated to produce the same number of child models by shifting the requisite hyperparameter by the mutation shift value (**Table 4**). The remaining 20 worst-performing (highest BIC) solutions were discarded. The next generation of models was then formed by adding 60 new models with randomized settings and feature sets and adding these to the 40 child models retained from the initial generation. Thereafter, IEL continued to recombine, mutate and discard 100 models per generation in a similar fashion to minimize the BIC until the latter fitness function plateaued. With 100 models fitted per generation, IEL typically fits ∼40,000 models per experiment over ∼400 generations.

IEL jointly performs this optimization across hyperparameter settings with automated feature selection to mitigate the risk of overfitting. During training, IEL has available to it the total set of features selected through the feature filtering process. To fit each model in the initial generation of 100 models, IEL selects a random number of features in the range [2-15] randomly samples specific features from the set of available features. After computing the BIC, feature sets from the best-performing 60 models were individually allocated to the recombined and mutated child models. Other feature sets were discarded. As with hyperparameter tuning, this process was repeated for succeeding generations until the BIC plateaued.

IEL implements recursive learning to promote computational efficiency. After training until the BIC plateaus, the elbow of the fitness function is determined by plotting fitness versus number of features and re-start learning with a warm start. The feature set available after this warm start is constrained to that subset of features, thresholded by their importance (**Feature importance determination**), corresponding to the elbow. Learning then proceeds by thresholding features available for learning at the original warm start feature importance + 2 standard deviations. In addition, the number of models per generation is reduced to 50, where 20 models are recombined and 10 models are mutated. Otherwise, training after the warm start proceeds as detailed above.

### Cross validation

During training, every one of the 100 models in each learning generation was trained and validated using cross-validation stratified *k*-fold cross validation. As detailed above, IEL allows the number of features used to fit each model to differ within each model in every generation. Accordingly, *k* (the number of splits) was set as the nearest integer above [sample size/number of features]. Cross validation was implemented with the scikit-learn StratifiedKFold function.

### Testing for generalization in unseen test data and performance measurement

After training was completed, the 100 best-performing models in the training phase identified by IEL were tested on the holdout, unseen test set by applying the requisite hyperparameter settings and selected predictors. The AUROC, accuracy, precision, recall and F1 were computed for test set models using standard Sci-Kit learn libraries. The threshold for prediction probability was 0.5 and ROC curves were generated for each experiment.

### Feature importance determination

We computed feature and predictor importances during the training and testing phases respectively with the Shapley Additive Explanations (SHAP) technique as instantiated in the SHAP toolbox (https://shap.readthedocs.io/en/latest/). SHAP is a game theoretic approach that may be used to explain the output of any ML model including ‘black box’ estimators such as artificial neural networks and is considered resistant to multicollinearity. (84) It unifies prior methods such as LIME, Shapley sampling values and Tree Interpreter. The SHAP algorithm is embedded inside IEL. Feature importances obtained during training are used to determine thresholds for recursive warm start learning. Predictor importances obtained during testing are used to quantify the relative importance of each predictor to obtaining the final predictions reported in **Results**. We computed SHAP scores at group- and person-levels. The former represent mean predictor importance across the participant sample and are the statistic typically reported in ML studies. The latter express predictor importance at the individual level and are important to a) calibrate the directionality of the relationship between predictor and target and to b) understand how predictor importance varies across the population of interest.

## RESULTS

All results presented below are from testing for generalization in the holdout, unseen test dataset.

### Predictive performance

Deep learning optimized with IEL was able to prospectively predict future, new onset cases of TD at 11-12 yrs using data collected at 9-10 yrs with high accuracy (92%), F1 (92%) and AUROC (0.96). All performance statistics may be viewed in **Table 5** with corresponding ROC curves in **Figure 2**. Of note, all performance statistics in predicting new onset cases of TD including recall and precision achieved >85%, a generally-accepted threshold for clinical utility.

**Table 5:** Performance of deep learning optimized with Integrated Evolutionary Learning in predicting cases of thought disorder in early adolescence. Performance statistics of accuracy, precision, recall, F1 and the AUROC are shown for the most accurate model obtained with deep learning optimized with Integrated Evolutionary Learning. We used features obtained at 9-10 years of age to predict new onset and all prevailing cases of TD at 11-12 years of age.

| Age of case determination | Accuracy (%) | Precision (%) | Recall (%) | F1 (%) | AUROC |
| --- | --- | --- | --- | --- | --- |
| New onset cases | 92.0 | 87.7 | 94.7 | 92.2 | 0.96 |
| All prevailing cases | 86.0 | 82.3 | 81.3 | 85.3 | 0.93 |

**Figure 2:**
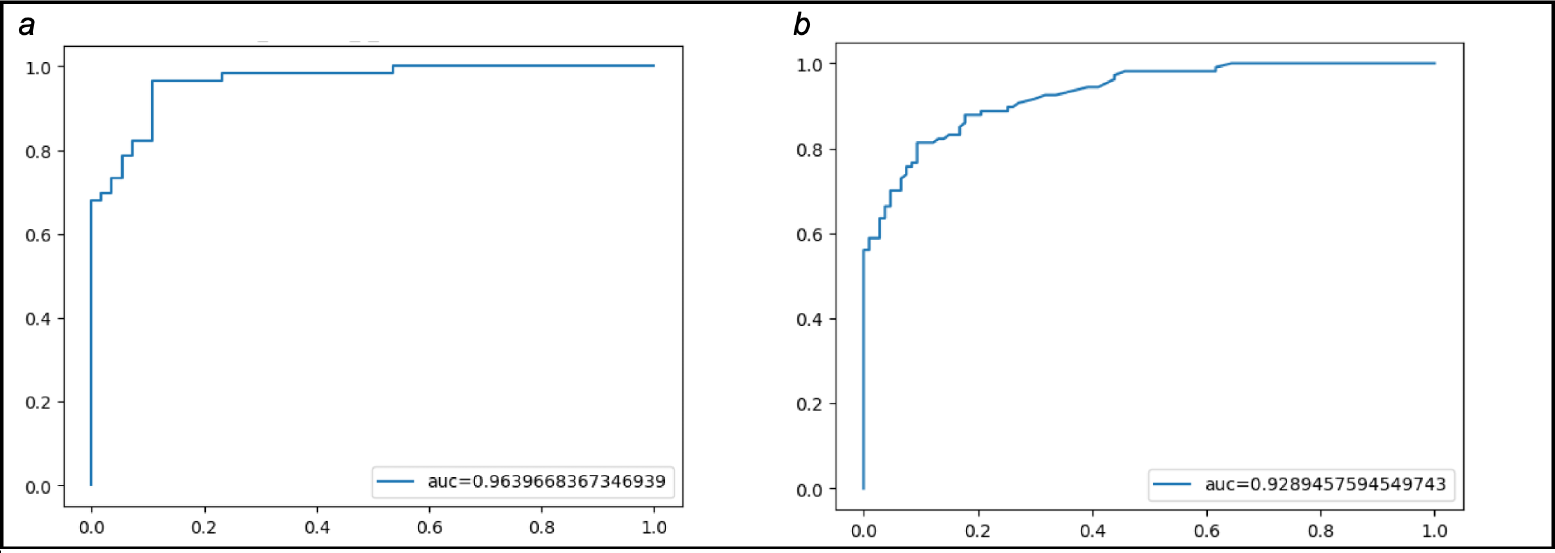
Receiver Operating Characteristic curves for deep learning optimized with Integrated Evolutionary Learning in predicting cases of thought disorder in early adolescence. Receiver Operating Characteristic (ROC) curves are displayed for the most accurate model predicting TD cases obtained with deep learning optimized with Integrated Evolutionary Learning for a) new onset cases; and b) all prevailing cases at 11-12 years of age. These ROC curves correspond to performance statistics shown in **Table 5**.

In our comparator analysis where all prevailing cases of TD at 11-12 yrs were prospectively predicted with features collected at 9-10 yrs, lower performance obtained. Accuracy reduced to 85.5% with a substantially lower F1 and AUROC at 84.7% and 0.93, respectively (**Table 5**).

### Predictors of thought disorder cases in early adolescence

Final predictors of TD cases were identified after optimization with IEL during the training process and testing in held-out, unseen data (**Table 6**) and the directionality of their relationship with case status determined by inspecting plots of person-level SHAP importances (**Figure 3**). In new onset TD cases, the most important predictors (quantified with mean SHAP scores) were the total sleep disturbance score based on the Sleep Disturbance Scale for Children (85) and whether the youth had come to clinical attention for mental health or substance use services prior to age 9-10. The youth’s prosocial behaviors score had an inverse relationship with

**Table 6:** Predictors of thought disorder in early adolescence. Optimized predictors of new onset TD cases at 11-12 years as well as all prevailing cases at 11-12 years of age are shown for the most accurate models obtained using deep learning optimized with Integrated Evolutionary Learning. The average importance of each feature to obtaining the prediction is computed with the Shapley Additive Explanations technique (SHAP). Features in blue indicate a negative relationship with TD discerned by mapping person-level SHAP importances (**Figure 3**). MH = mental health; SU=substance use; ROI = region of interest.

| Age of case determination | Ranked Final Predictors | Importance |
| --- | --- | --- |
| New onset at age 11-12 years | Total sleep disturbance score | 0.20 |
|  | Ever received MH/SU services | 0.14 |
|  | Prosocial behaviors score | 0.06 |
|  | Parent total behavioral problems score | 0.04 |
|  | Parent reports some conflict with youth | 0.04 |
|  | T1 intensity left putamen ROI | 0.03 |
|  | Parent externalizing syndrome score | 0.01 |
|  | Mean | 0.07 |
| All prevailing cases at age 11-12 years | Ever received MH/SU services | 0.19 |
|  | Parent total behavioral problems score | 0.15 |
|  | Prosocial behaviors score | 0.07 |
|  | Parent reports some conflict with youth | 0.03 |
|  | Mean | 0.11 |

**Figure 3:**
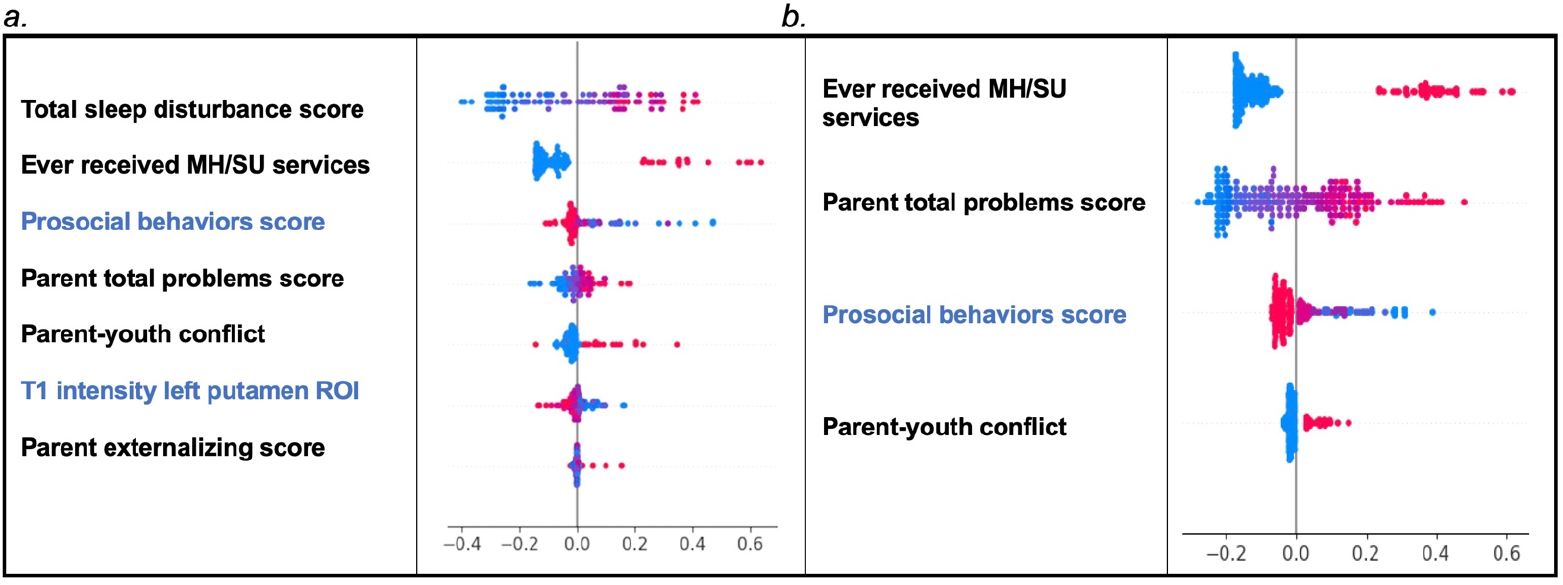
Person-level predictor importances of new onset and prevailing cases of Thought Disorder. Summary plots are presented of the importance of each final predictor (computed with the Shapley Additive Explanations technique) on an individual subject level to predicting a) TD with new onset at 11-12 yrs and b) all prevailing cases of TD at 11-12 yrs with features collected at 9-10 yrs. The color gradient represents the original value of each feature where red = high and blue = low. Discrete (binary) features appear as red or blue, while continuous features appear as a color gradient.

new onset case status. A collection of parent-related factors was also prominent: parental T scores for total behavioral problems and the externalizing subscale on the ABCL (the adult equivalent to the CBCL) and the degree of conflict between parent and child. While psychosocial predictors predominated in both our new onset and prevailing cases models, prospective prediction of new onset TD cases was notable for differences in left putamen structure appearing as a high-value final predictor. Specifically, average intensity of the normalized T1-weighted image within the left putamen region of interest as obtained with the FreeSurfer whole brain aseg segmentation. This neural metric had an inverse relationship with new onset case status.

Final, optimized predictors in our comparator model of all prevailing cases of TD at 11-12 yrs were a subset of those for new onset cases: whether the child had come to clinical attention prior to 9-10 yrs; parent total behavioral problems score on the ABCL; parent/child conflict; and the child’s prosocial behaviors score (inverse relationship). Thus, the new onset of TD at 11-12 yrs was specifically differentiated by the additional predictors of sleep disturbances, structural differences in the putamen and the level of parent externalizing behaviors obtaining when the child was 9-10 yrs.

### Person-level predictors of thought disorder in early adolescence

We computed person-level predictor importance (SHAP scores) across each participant sample to investigate the relationships between predictors and TD cases at the individual level. These may be viewed in SHAP summary graphs in **Figure 3**. Mapping of person-level SHAP importances reveals that features that are relatively more important to predicting case status in TD also tend to have more widely dispersed person-level importances i.e. greater inter-individual variability across the participant sample.

## DISCUSSION

The onset of new TD in early adolescence is of substantial research and clinical interest given its status as the strongest predictor of schizophrenia, where the latter tends to onset in late adolescence to early adulthood (slightly later for females) after a prodromal period. (86-89) While many studies have constructed prospective predictive models of the onset of schizophrenia, the onset of TD has been less studied. The emergence of large, naturalistic cohorts such as ABCD have opened new opportunities to explore the onset of thought problems and TD earlier in adolescence in unstratified populations. While the CBCL thought problems subscale was not explicitly designed to detect prodromal schizophrenia, it has demonstrated equivalent efficacy to the DSM-derived Psychotic Symptoms Scale (DOPSS) in detecting clinically-significant psychosis in youth. (75) This is notable, since the CBCL is a very widely-used instrument in child and adolescent health care, often as part of a core battery of routine assessment.

Here, we constructed prospective models in early adolescence and identified 7 factors obtaining at 9-10 yrs that predicted future TD onset at 11-12 yrs with very high accuracy (92%) and reliability (F1=92%; AUROC=0.96) after a large-scale optimization process. Given the paucity of prospective predictive modeling of TD, we cast a wide net and analyzed nearly 6,000 available features including 5,014 multimodal neural metrics. Of note, the final predictors we identified represent a set of non-linear interactions. Thus, our work suggests that the onset of TD in early adolescence is predicted by neural x psychosocial interactions. Specifically, between structural differences in the left putamen and factors pertaining to the parental environment (parent total behavioral problems and externalizing behaviors; conflict between parent and child) as well as non-specific behavioral issues in the child (reduced prosocial behaviors and sleep problems). Two comparisons are instructive. Our own and others’ work in the early waves of the ABCD cohort reveals that few of these factors taken individually are specific to TD in early adolescence -- other than structural abnormalities in the putamen. (90-94) Further, we here provide a comparator model predicting all prevailing cases of TD at 11-12 yrs that implicated 4 predictors that were a subset of the 7 factors that predicted new onset cases. These comparisons suggest that it is the particular set of interacting predictors and presence of putaminal structural differences in late childhood that is specific to TD onset in early adolescence.

The putamen is a subcortical deep brain nucleus that together with the globus pallidus forms the lentiform nucleus which in turn combines with the caudate nucleus to comprise the striatum and basal ganglia. The putamen is a dopamine-enriched structure involved in motor control and learning where volumetric differences have previously been associated with schizophrenia spectrum disorders and degenerative neuropsychiatric conditions. (95) For example, volumetric decreases in the left putamen and caudate are specifically associated with the presence of delusions in first episode schizophrenia (FEP). (96) The left putamen is also well-associated with cognitive (rather than motor) aspects of language processing, in particular phonological processing, articulation and language discrimination, including suggestive links to dopamine metabolism. (97-99) For instance, Tettamini et al used [11C]raclopride and PET to demonstrate a correlation between left putamen dopamine release and phonologic processing speed and error detection. (100) Moreover, the putamen is a known site of antipsychotic action and baseline activity in the left putamen specifically predicts response to olanzapine in drug-naïve, FEP patients. (101) Specific structural and functional neural correlates have been associated with TD in patients with schizophrenia, again primarily in language-related brain regions including cortical and subcortical areas. (102-106) However, studies in people without a diagnosis of schizophrenia, particularly youth, are lacking. (107) To our knowledge, ours is the first study that has identified the left putamen as a specific, prospective predictor of the new onset of TD in youth. Here, this brain ROI effectively competed against 5,013 other neural metrics and numerous other candidate predictors over ∼40,000 model fits for inclusion as a final and specific predictor of TD onset. In the context of prior evidence linking the left putamen to language processing, dopamine release and antipsychotic action, left putamen structure may therefore be advanced as a candidate biomarker of TD onset in early adolescence.

Other predictors of youth TD that we identified as specific to new onset cases were sleep disturbances and the level of parental externalizing behaviors. Sleep disturbances and sleep loss or insomnia are well known correlates of schizophrenia symptoms and illness severity, with many psychotic episodes preceded or even perhaps precipitated by sleep loss including both prodromal and relapsing patients. (108-111) In the present study, many different sleep-related features were available in the optimized selection process including different types of sleep disorders identified using the Sleep Disturbance Scale for Children. However, the total burden of sleep disturbances was selected as a final and specific predictor of TD onset, suggesting the nature of the sleep disturbance is less defined in TD than in other youth neuropsychiatric conditions. For example, ADHD tends to be associated specifically with longer sleep latency and excessive daytime somnolence. (112, 113)

In terms of parental-related factors, we found that the total burden of parent behavioral problems (as quantified by the ABCL) and conflict between parent and child were shared predictors of new onset and prevailing cases of TD, where the level of parent externalizing behaviors was specific to new onset cases. Adverse child rearing environment is a known risk factor for the development of schizophrenia and posited as contributing to gene x environmental interactions in disease etiology. Risk for schizophrenia onset has been associated with earlier parental loss, poor mothering and atypical mother-child interactions. (114-116) For instance, persons with schizophreniform disorder (but not other major psychiatric conditions) in the Dunedin longitudinal study were significantly more likely to have had atypical mother-child interactions. (117) Frank child abuse also appears to be a risk factor for the onset of schizophrenia, though it has been more strongly associated with positive symptoms rather than TD. (118) While we are not aware of studies that have specifically examined links between the child-rearing environment and TD onset separately from or preceding schizophrenia, our results suggest that the tenor of the relationship between parent and child, and increased externalizing and problem behaviors in parents may contribute to the likelihood of TD onset in early adolescence.

In conclusion, our work suggests that left putamen structural differences, sleep disturbances and the level of parent externalizing behaviors in late childhood are specific predictors of new onset TD in early adolescence and that interactions between these factors, low youth prosociality and other parent-youth related stressors (parent behavioral problems, parent-child conflict) are a specific risk constellation driving TD onset. Structural differences in the left putamen in late childhood represent a potential candidate biomarker of risk for TD (and perhaps later onset schizophrenia) warranting further investigation. Of note, an important predictor of TD cases at 11-12 yrs was whether a child had come to clinical attention at or before 9-10 yrs, suggesting that opportunities for screening and intervention may exist prior to the onset of frank TD. Moreover, our work suggests that extending current interventions for sleep promotion and psychological therapies developed for people with schizophrenia spectrum disorders to address disturbed sleep, psychosocial stressors, maladaptive family function and communication styles earlier in adolescence may be worthy of exploration to modulate risk for TD. (119-122)

## LIMITATIONS

The present study analyzes secondary data collected in the ABCD study. As such, we cannot control for any data collection bias. The ABCD study aimed for population representation, though there is currently a mild bias toward higher-income participant families of self-reported white race. Here, cases were matched with controls based on natal sex and age. However, sex, gender, race, ethnicity and many sociodemographic and cultural factors were included as candidate predictors as is standard practice in large scale ML studies and which does allow for the influence of such factors to be revealed. Further work using a similar design in participant samples stratified by sex/gender, race/ethnicity or other demographic factors could elucidate differential results. Data is not available prior to baseline (age 9-10 years) assessment and we cannot therefore conclusively rule out that youth participants may have met criteria based on the CBCL for TD prior to ≤8 yrs. Thus, it is possible that certain cases coded as ‘new onset’ at 11-12 years of age could have met clinical criteria at ≤8 yrs but not at 9-10 yrs. In the present study, we defined cases as an individual meeting ASEBA clinical thresholds in the CBCL Thought Problems subscale score and did not exclude participants who were above clinical thresholds in other subscales. Thus, co-morbidity may be present in the experimental samples as is common in clinical populations and occurs in most research studies in early adolescence. Our study is not exhaustive. It is possible that different results could have been obtained if more or different candidate predictors were included. We tested for generalization in a holdout, unseen test set obtained by partitioning the data, a gold standard method in ML. However, methods and results should also be tested for replication in an external dataset other than ABCD.

## Data Availability

All data produced in the present work are available upon reasonable request to the authors

https://nda.nih.gov/abcd/abcd-annual-releases.html

## FUNDING

Research reported in this publication was supported by the National Institute of Mental Health of the National Institutes of Health under award number **R00MH118359** to NdL. The content is solely the responsibility of the authors and does not necessarily represent the official views of the National Institutes of Health.

## ACKNOWLEDGEMENTS

Data used in the preparation of this article were obtained from the Adolescent Brain Cognitive Development^SM^ (ABCD) Study (https://abcdstudy.org), held in the NIMH Data Archive (NDA). This is a multisite, longitudinal study designed to recruit more than 10,000 children age 9-10 and follow them over 10 years into early adulthood. The ABCD Study® is supported by the National Institutes of Health and additional federal partners under award numbers U01DA041048, U01DA050989, U01DA051016, U01DA041022, U01DA051018, U01DA051037, U01DA050987, U01DA041174, U01DA041106, U01DA041117, U01DA041028, U01DA041134, U01DA050988, U01DA051039, U01DA041156, U01DA041025, U01DA041120, U01DA051038, U01DA041148, U01DA041093, U01DA041089, U24DA041123, U24DA041147. Additional support for this work was made possible from NIEHS R01-ES032295 and R01-ES031074. A full list of supporters is available at https://abcdstudy.org/federal-partners.html. A listing of participating sites and a complete listing of the study investigators can be found at https://abcdstudy.org/consortium_members/. ABCD consortium investigators designed and implemented the study and/or provided data but did not necessarily participate in the analysis or writing of this report. This manuscript reflects the views of the authors and may not reflect the opinions or views of the NIH or ABCD consortium investigators. The ABCD data repository grows and changes over time. The ABCD data used in this report came from 10.15154/1523041. DOIs can be found at https://nda.nih.gov/abcd/abcd-annual-releases.html. The support and resources from the Center for High Performance Computing at the University of Utah are also gratefully acknowledged.

## Notes

### Competing Interest Statement

The authors have declared no competing interest.

### Author Declarations

The IRB of the University of Utah waived ethical approval for this work

